# 3D Printed Snorkel Mask Adapter for Failed N95 Fit Tests and PPE Shortages

**DOI:** 10.1101/2020.08.18.20174540

**Authors:** Shiv Dalla, Rohit Shinde, Jack Ayres, Stephen Waller, Jay Nachtigal

**Affiliations:** University of Kansas Medical Center; Unaffiliated

## Abstract

**Introduction:** The shortage of personal protective equipment (PPE) across the country has been widely discussed throughout the COVID-19 pandemic. Unfortunately, recent reports indicate that PPE shortages persist amidst continually increasing caseloads nationwide. Additionally, there have been reports of poor-fitting masks, a problem which is magnified by shortages. The lack of adequate access to conventional N95 masks pushed for some to pursue 3D printing and locally distributing their own manufactured masks as substitutes when PPE, including N95 masks, were not readily available. The design presented, the snorkel mask adapter, is one such design born from the local maker community in partnership with local physicians and hospitals. This article discusses the design, manufacturing, and validation of the snorkel mask adapter and its immediate use in the COVID-19 pandemic as well as future use as stopgap PPE.

**Methods:** The design presented is an adapter which can be used with a commercially available snorkel mask in order to serve as a full face respirator in either the case of a PPE shortage or more pertinently for those who are unable to pass fit testing with the available N95 respirators at their respective facilities. Mask components were 3D printed, assembled, and then fit tested by qualitative fit testing (QLFT) at The University of Kansas Health System (TUKHS) in Kansas City, KS as a proof of concept.

**Results:** At TUKHS, the mask was fit tested on 22 individuals who required an N95 mask but were not able to pass qualitative fit testing with the masks available to them at the time. Of the 22 tested, all 22 of them were able to pass QLFT with the snorkel mask, adapter, and viral/bacterial filter combination.

**Conclusion:** The results of the fit testing at TUKHS is promising for this N95 alternative. More extensive testing can and should be done, including quantitative fit testing. Persistently increasing caseloads and PPE shortages necessitates an urgent dissemination of these preliminary results. The authors do not advocate for this design as a replacement of traditional N95 masks or other PPE but do endorse this design as a stopgap measure, proven to be effective in situations of dire PPE shortage or for individuals who have failed fit testing with conventional PPE.

## INTRODUCTION

### Background

In December of 2019 the first cases of a novel coronavirus, named SARS-CoV-2, were reported in China.^1^ As the virus spread across the continent, it proved to be both highly contagious and highly virulent.^1^ Some infected patients were suffering from acute respiratory distress syndrome (ARDS).^1^ By the middle of March 2020, the World Health Organization had designated the COVID-19 outbreak as a pandemic. Reports of not only strained economies but also strained global healthcare systems flooded the news.^2^ Within healthcare settings, the focus shifted to the adequacy of personal protective equipment (PPE).^3^

### PPE Shortages

The shortage of PPE across the country has been widely discussed by government officials and healthcare practitioners as well as the general population.^3-5^ Unfortunately, recent reports indicate the supply of PPE “is running low again as the virus resumes its rapid spread and the number of hospitalized patients climbs.”^6^ Although the situation has improved since it was first evaluated earlier in the year, many experts predict that shortages persist.^7^ Additionally, some predict that it may take years in order for stockpiles to fully replenish, an increasingly urgent challenge as cases continue to rise across the United States.^7^ Shortages of PPE have repeatedly been shown to contribute to the spread of the virus, and “the situation is especially dire at hospitals serving communities of color or patients on Medicaid.”^8^

### Fit Testing

In addition to PPE shortages, various studies have commented on difficulties regarding inadequate mask fitting.^9,10^ Mask fit is an important consideration of mask efficacy and is typically measured by either qualitative fit-tests (QLFT) or quantitative fit-tests (QNFT).^9^ In qualitative fit-tests, mask wearers are tested to see if they can detect bitter or sweet scents aerosolized around the masked wearer, whereas quantitative fit-tests measure ratios of ambient aerosols inside and outside of the mask.^9^ A 2018 Korean study reported QNFT pass rates of the four most common N95 models to be below 50%, with poorer fit test results observed for women compared to men.^10^ The inherent challenge of the possibility of a poor mask fit is magnified during pandemic settings due to time pressures and limited availability of options.

### 3D Printed Alternatives

As the aforementioned issues with PPE were playing out, communities of 3D printing aficionados, colloquially known as “makers,” began to discuss the issue and created homemade masks, face shields, and gowns. Specifically, those with access to 3D printers were encouraged to continually run their machines to produce face-shields and masks that could be used in the health care sector. The lack of adequate access to conventional N95 masks pushed for some to pursue 3D print and locally distribute masks to assist in reducing PPE shortages. The widespread availability of 3D printers and cost-effective nature of the manufacturing process makes it excellent for such purposes.

The design presented is an adapter which can be used with a commercially available snorkel mask in order to serve as a full face respirator in either the case of a PPE shortage, or more pertinently for those who are unable to pass fit testing with the available N95 respirators at their respective facilities. This article focuses on the design, manufacture, and validation of the snorkel mask adapter design and its usage in the COVID-19 pandemic as well as future usage as stopgap PPE.

## METHODS

### Adapter Design

The adapter was designed to connect an OceanReef snorkel mask to a Hudson RCI In-Line Bacterial/Viral Filter^11^ commonly found in both operating rooms and intensive care units for filtration and humidification of air in mechanically ventilated patients. These filters were chosen for their cost-effective nature, high availability during PPE shortages, and low resistance to gas flow while maintaining 99.99% filtration efficiency.^11^ The assembled design is shown in Figure 1 and the 3D printed adapter is shown in Figure 2.

**Figure 1:**
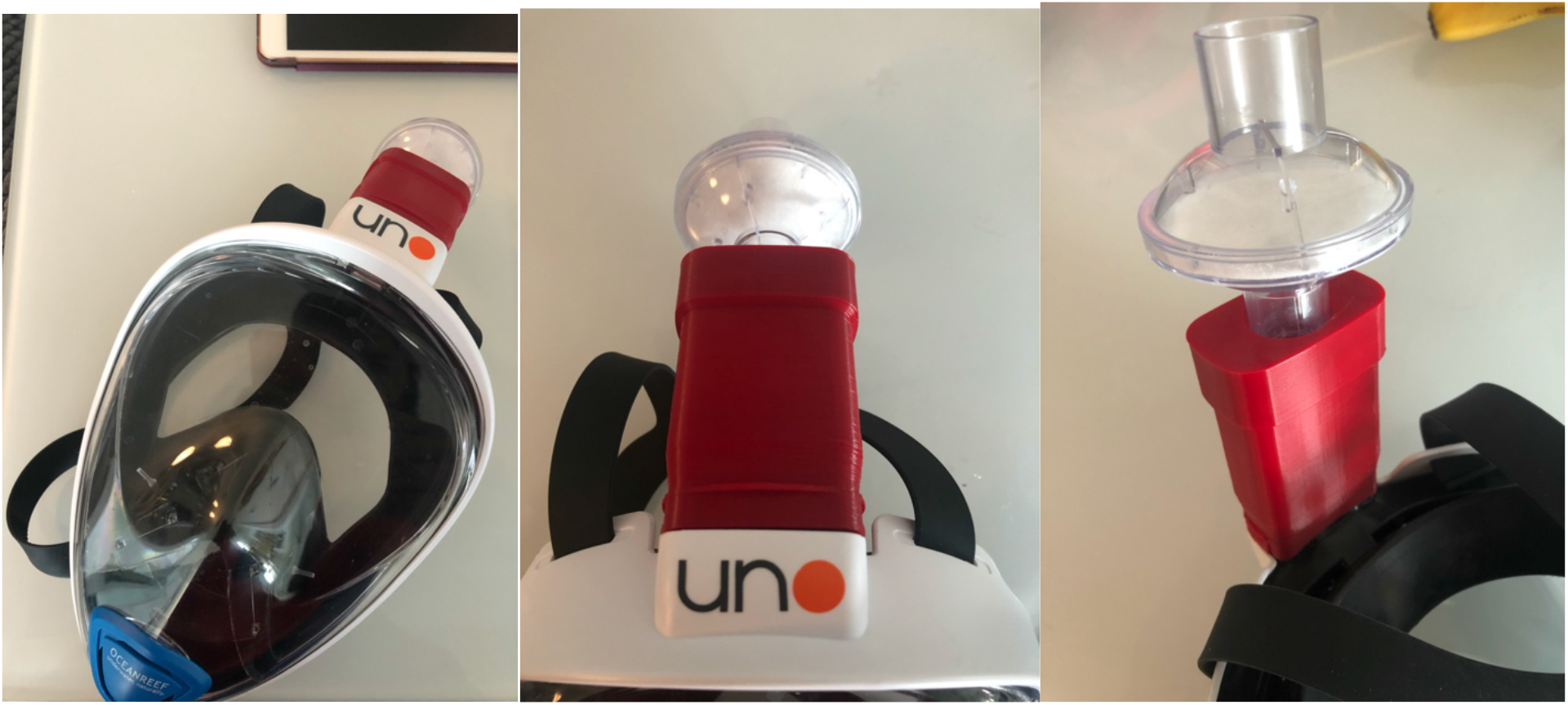
OceanReef snorkel mask coupled to a Hudson bacterial/viral filter with the 3D printed adapter (red)

**Figure 2:**
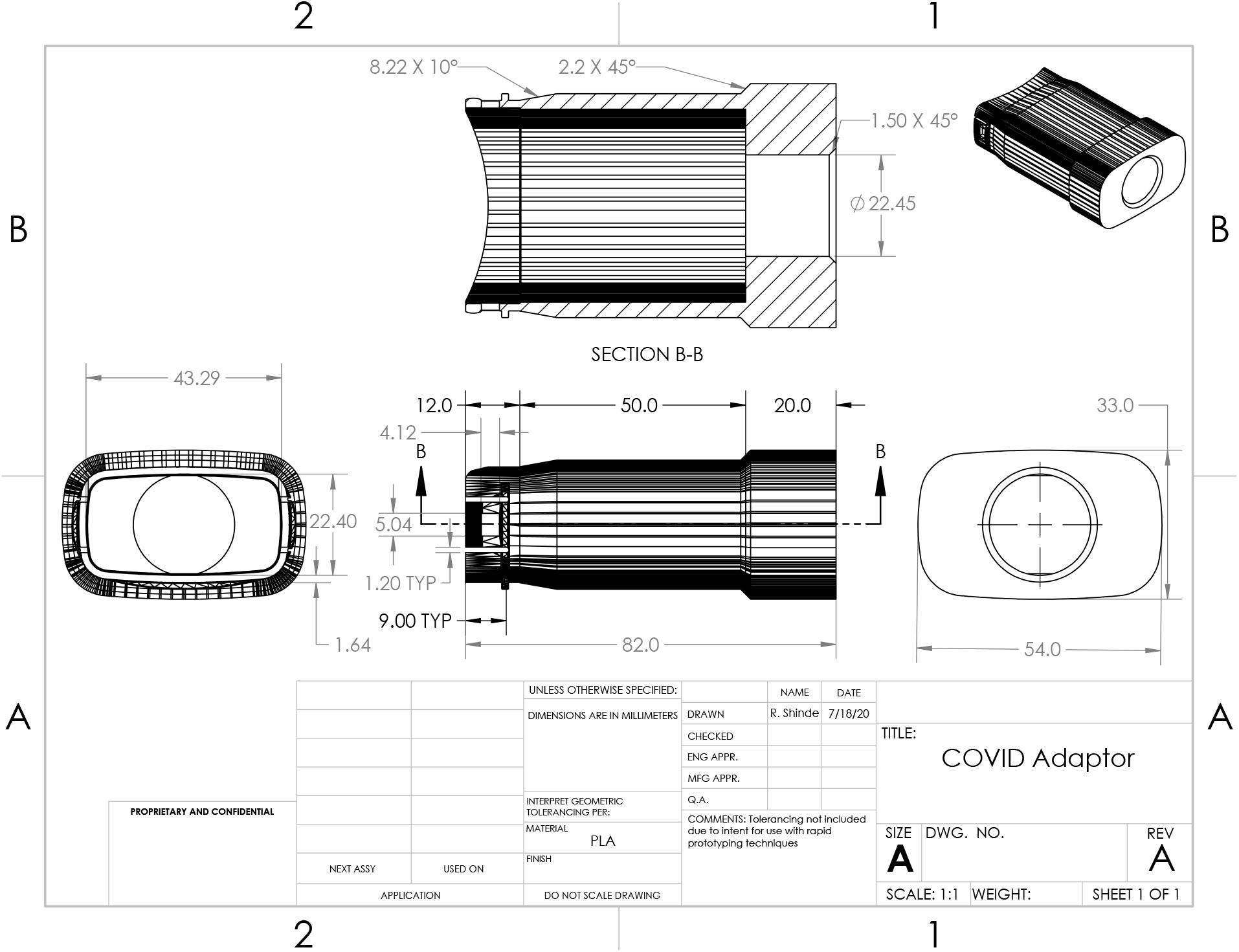
Engineering Drawing of Snorkel Mask Adapter Design.

### Design and 3D Printing

This design was generated using commercial CAD software and exported to an Standard Tessellation Language (STL) file for compatibility with hobbyist 3D printer workflows. These workflows use a “slicer” software to convert the universal STL file into GCODE that is specific to an individual 3D printer. Two combinations of slicer and 3D printer were tested: the Cura slicer was used to prepare GCODE for a Lulzbot Taz5, and PrusaSlicer was used to prepare GCODE for a Creality CR-10S. Adapters were printed in a variety of filaments including Polylactic acid (PLA), Polyethylene terephthalate glycol (PET-G), and Thermoplastic polyurethane (TPU). Polylactic acid (PLA) was chosen given its melting point, ease of use, and cost effectiveness. Print settings shown in Table 1 were used. However, these may vary based on software and printer. For most printers, the default PLA settings will be sufficient.

**Table 1:** Print settings used on the Taz5 Printer with eSun PLA+ Filament.

| Select Print Setting | Value |
| --- | --- |
| Layer Height | 0.25mm |
| Wall Line Count | 4 |
| Top/Bottom Layers | 4 |
| Infill Density | 20% to 50% |
| Printing Temperature (Nozzle) | 205°C |
| Printing Temperature (Build Plate) | 60°C |

### Usage and Fit Testing

Before being taken in for formal qualitative fit testing (QLFT) at The University of Kansas Health System (TUKHS), a water seal test was performed. The wearer donned the mask, adapter, and viral filter combination and was submerged into water such that the water level was above the connection of the filter to the adapter, but below the top of the filter. This was done to allow for identification of leaks in the system. If the wearer felt water entering the mask or if air was seen escaping in the adapter fitment the model was identified as not air or water tight. This test was only performed before a version of the adapter was sent for QLFT at TUKHS.

At TUKHS, the design was fit tested by the facilities staff using a standard saccharin solution aerosol protocol. At TUKHS, participants were those who were not able to pass fit testing with the standard N95 respirators available at TUKHS. None of the designers of the adapter or authors of this paper were present for fit testing, to ensure an unbiased fit test.

### Study Design

IRB approval was not requested nor required for this study because this was essentially a proof of concept and quality improvement initiative. The design went through the proper channels at TUKHS for testing and approval of PPE during the height of the pandemic. Although further analysis and study to prove efficacy is required, the purpose of this study and article is to discuss the design, manufacturing, and validation of the snorkel mask adapter concept.

## RESULTS

At TUKHS, 3832 people have been fit tested at least once for standard half face respirators. Of these, 211 people are currently listed as powered air purifying respirator (PAPR) only, placing a significant load on access to PAPR devices. The snorkel mask, adapter, and viral filter combination was fit tested on 22 individuals of the 3832 people who required an N95 mask but were not able to pass qualitative fit testing with the masks available to them at the time. Three sizes of Ocean Reef Uno snorkel masks were used: S/M, M/L, L/XL. Of the 22 individuals tested, all 22 of them were able to pass QLFT with the snorkel mask (one of 3 sizes), adapter, and Hudson RCI viral/bacterial filter combination. It was deemed appropriate that five of these 22 individuals were given snorkel masks, adapters, and filters to use as necessary.

## DISCUSSION

The results of the fit testing at TUKHS is promising for this N95 alternative. More extensive testing can and should be done, including quantitative fit testing. However, the initial results suggest that this mask could be efficacious at a larger scale. Of those who passed qualitative fit testing (QLFT) (N = 463) with all N95 FFR models, 86.9% (N = 459) also passed quantitative fit testing. This suggest that qualitative fit testing, although less rigorous than quantitative fit testing, does correlate highly with proper mask fitment.^9^

Persistently increasing caseloads and PPE shortages necessitates an urgent dissemination of these preliminary results. The authors do not advocate for this design as a replacement of traditional N95 masks or other PPE but do endorse this design as a stopgap measure, proven to be effective in situations of dire PPE shortage or for individuals who have failed fit testing with conventional PPE.

## CONCLUSION

The results of the fit testing at TUKHS is promising for this N95 alternative. More extensive testing can and should be done, including quantitative fit testing. Persistently increasing caseloads and PPE shortages necessitates an urgent dissemination of these preliminary results. The authors do not advocate for this design as a replacement of traditional N95 masks or other PPE but do endorse this design as a stopgap measure, proven to be effective in situations of dire PPE shortage or for individuals who have failed fit testing with conventional PPE.

## Data Availability

Any supplementary materials are listed in the references section of our manuscript.

## REFERENCES

1. Sohrabi C, Alsafi Z, O’Neill N, et al. World Health Organization declares global emergency: A review of the 2019 novel coronavirus (COVID-19). Int J Surg. 2020;76:71-76.

2. Mike-Hana Fongang M, Ahmadi N. The Impact of a Pandemic (COVID-19) on the Stock Markets: A Study on the Stock Markets of China, US and UK [Student thesis]2020.

3. Jacobs A, Richtel M, Baker M. ‘At War With No Ammo’: Doctors Say Shortage of Protective Gear Is Dire. The New York Times. 2020/03/19/T15:58:44-04:00, 2020;Health.

4. Jessop ZM, Dobbs TD, Ali SR, et al. Personal Protective Equipment (PPE) for Surgeons during COVID-19 Pandemic: A Systematic Review of Availability, Usage, and Rationing. Br J Surg. 2020.

5. Mandrola J. [CoViD-19 and PPE: some of us will die because of the shortage.]. Recenti Prog Med. 2020;111(4):183.

6. Fassett GM, Camille. Supply of PPE for medical workers begins to run low again as COVID-19 spikes in US. chicagotribunecom. 2020.

7. Pulver D, Wedell K, Mansfield E. Despite warnings, the US wasn’t prepared with masks for coronavirus. Now it’s too late. USA TODAY. 2020.

8. Dunn L, Fitzpatrick S. Few N95 masks, reused gowns: Dire PPE shortages reveal COVID-19’s racial divide. NBC News. 2020.

9. Hon CY, Danyluk Q, Bryce E, et al. Comparison of qualitative and quantitative fit-testing results for three commonly used respirators in the healthcare sector. J Occup Environ Hyg. 2017;14(3):175-179.

10. Huh YJ, Jeong HM, Lim J, et al. Fit Characteristics of N95 Filtering Facepiece Respirators and the Accuracy of the User Seal Check among Koreans. Infect Control Hosp Epidemiol. 2018;39(1):104-107.

11. Teleflex. Hudson RCI In-Line Bacterial Viral Filter For CPAP/BIPAP Machines. https://www.healthproductsforvou.com/p-hudson-rci-in-line-bacterial-viral-filter-for-cpapbipap-machines.html. Published 2020. Accessed 7-23, 2020.

